# Treatment Outcomes of Doxycycline Use for Musculoskeletal Infections: A Systematic Review and Meta-Analysis

**DOI:** 10.1101/2024.03.12.24304130

**Authors:** Rawabi Aljadani, Carolina Gonzalez Bravo, Matida Bojang, Martha L. Carvour

**Affiliations:** University of Iowa College of Public Health, Iowa City, Iowa, United States; University of Iowa Carver College of Medicine, Iowa City, Iowa, United States; Medical College of Wisconsin, Milwaukee, Wisconsin, United States

**Keywords:** doxycycline, musculoskeletal infections, treatment outcomes, systematic review

## Abstract

Doxycycline is among the most commonly used antibiotics for the treatment and long-term suppression of musculoskeletal infections such as osteomyelitis and periprosthetic joint infection. We systematically reviewed clinical trials and cohort studies that examined outcomes of musculoskeletal infections treated with doxycycline. Eligible studies were published in Medline or Embase in English before March 2, 2021. Eleven reports were included; eight addressed medical/non-dental infections, and three addressed dental infections. *Brucella* was the most frequently studied organism in the non-dental studies. Random-effects meta-analyses showed no significant difference in *Brucella* relapse risk after six weeks of treatment with 200 mg doxycycline compared to 400 mg ofloxacin daily (pool risk ratio: 0.94, 95% confidence interval: 0.2 - 4.45, I^2^ =L0%). Despite a large number of case reports, case series, and cross-sectional studies on this topic, few studies investigated doxycycline treatment outcomes; and evidence was largely limited to rare infections such as *Brucella*.

## INTRODUCTION

Musculoskeletal infections encompass a wide range of medical and dental conditions, including osteomyelitis, septic arthritis, pyomyositis, myositis, severe periodontitis, and periprosthetic joint infection. In clinical practice, confirming the diagnosis of musculoskeletal infection—including the microbiological cause and mechanism of infection—can be challenging (1, 2). Healthcare providers often use a combination of radiological imaging, laboratory testing, patient historical details (including key environmental exposures to potential pathogens), and clinical examination techniques to diagnose the infection and identify a treatment plan (3–5). Despite these combined efforts, many musculoskeletal infections do not have a known microbiological cause (35.4% - 72%) and are treated empirically, based on the organisms deemed most likely to be present (6–9). Such empiric treatment often requires antibiotic coverage for *Staphylococcus aureus,* the most common causative agent in musculoskeletal infections where an organism is identified (∼51%) (1, 10).

Doxycycline, a tetracycline antibiotic, is among the most commonly used antibiotics for the treatment—or long-term suppression—of musculoskeletal infections. Doxycycline has a broad range of activity against many Gram-positive bacteria, including *Staphylococcus aureus;* some Gram-negative bacteria; and some atypical bacteria, such as *Brucella, Borrelia,* or *Chlamydia*, making it an appealing choice for empiric regimens (11). Doxycycline also has a higher barrier to antimicrobial resistance compared to many other antibiotics, allowing for more reliable, sustained antibiotic effects over long courses of therapy, including prolonged courses used to treat many musculoskeletal infections (e.g., 1-2 months) and for chronic antibiotic suppression of musculoskeletal infections (e.g., months or years) (12–14). Furthermore, doxycycline has good bone penetration and oral bioavailability, placing it among the first-line choices when oral antibiotic agents are recommended for the treatment of a musculoskeletal infection (15, 16). Doxycycline is generally well tolerated, although it can cause some undesirable side effects including teeth discoloration due to its accumulation in the bone, photosensitivity, and gastrointestinal symptoms (11, 17). These combined characteristics of doxycyline—namely, its broad antibiotic spectrum of coverage, good oral bioavailability, and favorable side effect profiles—have led to increased reliance on oral doxycycline in antibiotic treatment regimens for musculoskeletal infections in recent years. In this study, we sought to systematically examine the existing evidence base for this evolving practice.

## METHODS

### Search strategy and articles selection

We systematically searched Medline and Embase for clinical trials or cohort studies investigating the outcome of treating musculoskeletal infections with doxycycline. We used the following search terms: (((((((Bone OR (Joint)) OR (Prosthetic joint)) OR (Osteomyelitis)) OR (Myositis)) OR (Pyomyositis)) OR (Septic Arthritis)) AND (Doxycycline)). Our search was restricted to include only studies published in English, studies conducted on humans, and studies published before March 2, 2021. The Medline search was conducted in March 2021; the Embase search was conducted in September 2021 using the same eligibility criteria. We excluded studies reporting only non-infectious syndromes, antimicrobial prophylaxis, sub-antimicrobial doses, or chronic antimicrobial suppression and studies that did not specifically assess or report musculoskeletal involvement of infections. Additionally, we searched the references cited within the included studies to identify any related records that were not captured by the original queries. Two researchers (R.A. and M.C.) independently screened the titles and abstracts for inclusion. For eligible studies identified during the screening process, the full texts were retrieved by R.A. and independently assessed for inclusion by R.A, C.G.B., and M.B. Any disagreements were resolved through discussion.

### Data extraction and quality assessment

Researchers R.A, M.B, and C.G.B. extracted the following data from all included articles: sample size, study population, site of infection, organism, doxycycline dose and duration, other antibiotics given in a combination with doxycycline, control or comparator dose and duration, follow up after treatment, and treatment outcome. Clinical trials were also subclassified as either medical/non-dental or dental trials to permit better interpretation of results. The quality of the included clinical trials was assessed independently by R.A, M.B, and C.G.B. using the Cochrane risk of bias tool – a two-part tool that evaluates the risk of selection, reporting, and other biases in the first part and the risk of performance, detection, and attrition bias in the second part and that classifies the risk of bias as high, low, or unclear (18). The quality of the cohort studies was examined using the Newcastle - Ottawa quality assessment scale for cohort studies, a star-based system that evaluates quality across three domains: selection, comparability, and outcome ascertainment (19).

### Data synthesis and statistical analysis

For clinical trials involving medical/non-dental outcomes and evaluating treatment relapse rates, we used the number of events and total number of subjects per treatment arm to compute the risk ratio (RR) with 95% confidence intervals (CIs) using a random-effects meta-analysis model. Additionally, for dental clinical trials, we used the reported means, standard deviations (SD), and total number of subjects per treatment arm to calculate the mean difference and 95% CIs using a random-effects meta-analysis model. We computed SDs for the means using the Cochrane Handbook for Systematic Reviews method, if a study reported the standard error of the mean (SEM) instead of an SD (20). We used *I*^2^ statistics to assess the degree of heterogeneity between the studies; however, because the number of included studies in the meta-analyses is small, we decided to use a random effect to construct all the meta-analyses in this study. Similarly, we did not generate funnel plots to assess publication bias, since the number of studies in the computed meta-analyses was less than 10 (20). RevMan 5.4 software was used to analyze the data and generate plots (20).

## RESULTS

### Search Results

We identified a total of 1135 reports published in Medline and Embase, and an additional 20 reports were found through citation searches. The most common types of published studies were case reports and case series (47.6%). After screening, 89 studies were referred for full-text review; and only 11 studies were eligible for inclusion after full-text review was completed. Of these reports, three were cohort studies (21–23), and eight were clinical trials (24–31). A flow diagram of the study identification, screening, and inclusion processes is shown in Figure 1.

**Figure 1.**
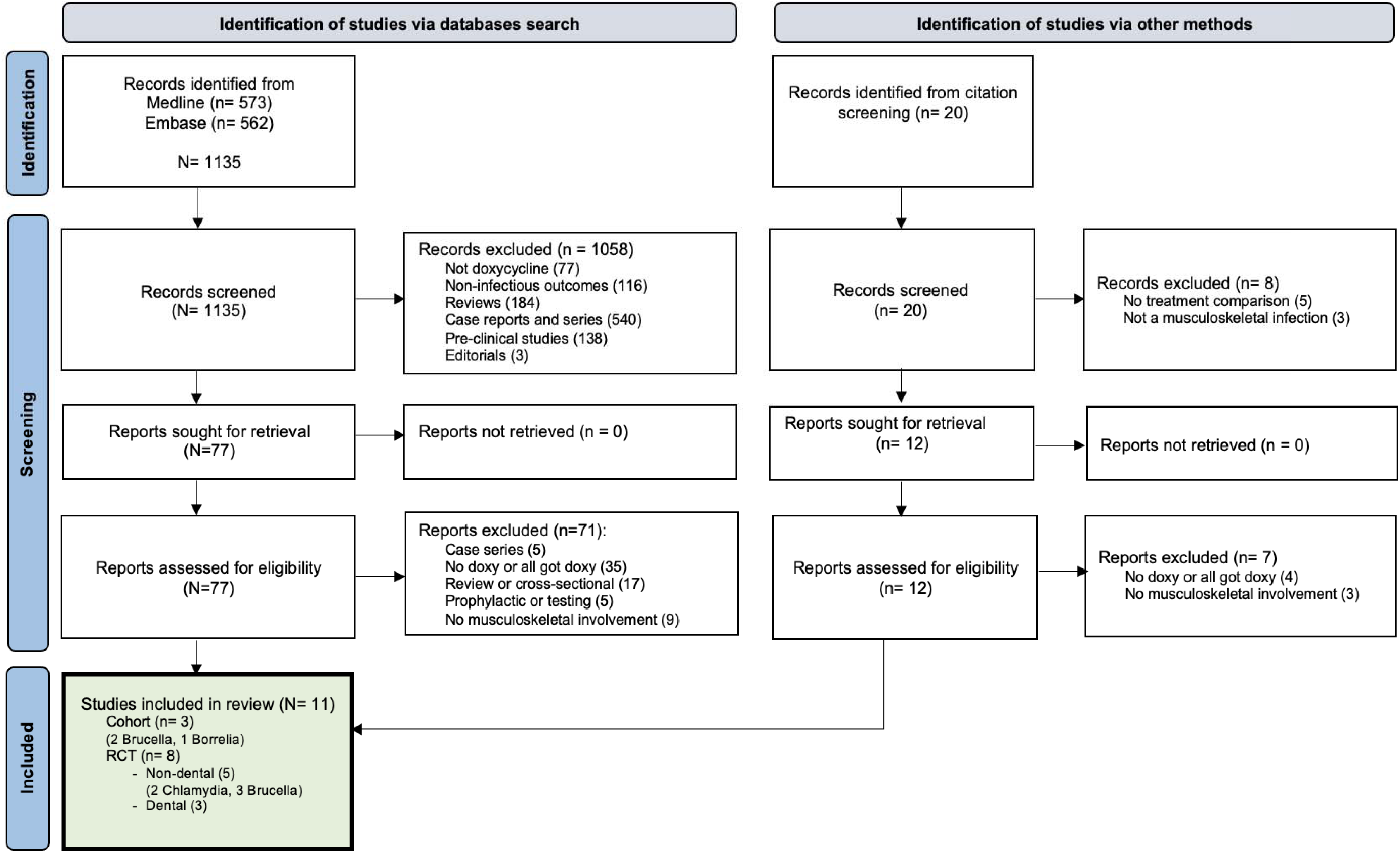
Flow diagram showing identification, screening, and inclusion of studies for systematic review.

### Site of Infection and Organisms

Among medical/non-dental musculoskeletal infections (Tables 1 & 2), *Brucella* (as a cause of arthritis and/or sacroiliitis) was the most investigated organism in both cohort studies and clinical trials (22, 23, 26–28), followed by *Chlamydia*-induced reactive arthritis (24, 25), and *Borrelia* (Lyme arthritis) (21). Dental studies mainly focused on periodontitis with bone involvement, regardless of the causative organisms (29–31).

**Table 1.** Summary of cohort studies eligible for inclusion in systematic review.

| Study | Country | Design | Sample size (N) | Study population | Infection site / Organism | Intervention |  | Comparison group | Follow up | Outcome |
| --- | --- | --- | --- | --- | --- | --- | --- | --- | --- | --- |
|  |  |  |  |  |  | Doxycycline | Additional antibiotics |  |  |  |
| **Grillon 2019** <sup>(18)</sup> | France | Retrospective | 37;<br>intervention:24;<br>comparison:13 | Adults | Arthritis / *Borrelia* | Doxycycline ; dose: NA; median duration: 4 weeks (range 3-12) | With or without ceftriaxone | Ceftriaxone, amoxicillin, tetracycline, or oral cefaclor | 17 months (range: 1-73) | Persistent synovitis: 34% (n=12); doxycycline 83% (10), ceftriaxone 33% (4), amoxicillin 8% (1). |
| **Hadadi 2014** <sup>(19)</sup> | Iran | Retrospective | 415;<br>intervention:170;<br>comparison:245 | Pediatric patients and adults (12 - 80 years) | Arthritis Sacroiliitis Spondylitis / *Brucella* | Doxycycline ; dose: NA; duration: NA | Rifampin or co-trimoxazole or both | Rifampin + co-trimoxazole, or rifampin + co-trimoxazole + gentamicin | Median follow-up: 7 months; maximum follow-up: 24 months | Relapse: 40 patients; intervention relapse rate: 2.5% (95% CI: 0-12.5%). Comparison relapse rate: 13.8% (95% CI: 8.2-20.5%) and 10% (95% CI: 5.1-16.3%) |
| **Fallatah 2005** <sup>(20)</sup> | Saudi Arabia | Retrospective | 159;<br>intervention:120;<br>comparison:20 | Pediatric patients and adults (13 - 40 years) | Joint involved / *Brucella* | Doxycycline ; dose: NA; duration: 3-4 weeks | Rifampin or streptomycin or co-trimoxazole | Rifampin + streptomycin or rifampin + co-trimoxazole | 4-16 weeks | Intervention relapse rate: 2.5%. Comparison relapse rate: 13.8% |

NA: not available or unknown.

**Table 2.** Summary of medical/non-dental clinical trials eligible for inclusion in systematic review.

| Study | Country | Design | Sample size (N) | Study population | Infection site / Organism | Intervention |  | Control | Follow up | Outcome |
| --- | --- | --- | --- | --- | --- | --- | --- | --- | --- | --- |
|  |  |  |  |  |  | Doxycycline | Additional antibiotics |  |  |  |
| **Putschky 2006** <sup>(21)</sup> | Germany | Randomized double-blind placebo-controlled trial | 37; intervention:15; control:14; withdrawal / loss to follow-up:5 | Adults, older than 18 (18–65) | Arthritis / *Chlamydia trachomatis* | 100 mg PO bid for 4 months |  | Placebo: one cap po bid for 4 months | 4 months | Mean differences: joint swelling: -1.5 (-2.8 to -0.2); joint tenderness: -2.3 (-3.8 to -0.8); erythrocyte sedimentation rate (ESR): -7 (-19 to 5); C-reactive protein (CRP): -1.4 (-3.5 to 0.7); pain intensity: -1.4 (-3.2 to 0.4) |
| **Smieja 2001** <sup>(22)</sup> | Canada | Randomized, triple-blind, controlled clinical trial | 60; intervention:30; control:30 | Adults, older than 18 (18–65) | Arthritis / *Chlamydia trachomatis* | 100 mg PO bid for 90 days |  | Identical-appearing placebo for 90 days | 90 days | Pain change scores: mean difference 1.5, (95% CI -1.2 to 4.2, p=0.25); composite functional change scores: mean difference 0.8 (95% CI -5.6 to 7.1, p=0.81) |
| **Alp 2006** <sup>(23)</sup> | Turkey | Controlled, non-randomized, open-label trial | 31; intervention:15; control:16 | 16 years and older | Spine / *Brucella* | 100 mg PO bid for at least 12 weeks | Streptomycin 1 g daily IM for 21 days | Ciprofloxacin 500 mg PO bid + rifampin 600 mg PO daily for at least 12 weeks | 12 months | Failure: 0 for both groups<br>Relapse: 0 for both groups |
| **Saltoglu 2002** <sup>(24)</sup> | Turkey | Clinical trial | 57; intervention:30; control:27 | 15-65 years | Arthritis Sacroiliitis / *Brucella* | 100 mg PO bid for 45 days | Rifampin 600 mg daily | Ofloxacin 200mg bid + rifampin 600mg daily | 6 months | Doxycycline/rifampin relapse rate: 6.7%; ofloxacin/rifampin relapse rate: 7.2% (P>0.05) |
| **Akova 1993** <sup>(25)</sup> | Turkey | Randomized clinical trial | 61; intervention:30; control:31 | Adults, older than 18 | Spondylitis, hip and knee arthritis, and sacroiliitis / *Brucella* | 200 mg PO daily for 6 weeks | Rifampin 600 mg daily | Ofloxacin 400 mg + rifampin 600 mg daily for 6 weeks | Monthly (3 months) and every 3 months (year) | Failure: intervention: 0; control: 1<br>Relapse: intervention: 1; control: 1 |

PO: orally; bid: twice a day; IM: Intramuscular injection.

### Interventions and Comparisons

*Brucella* studies compared doxycycline in combination with rifampin (22, 23, 27, 28), streptomycin (23, 26), or co-trimoxazole (22, 23) versus rifampin in combination with streptomycin (23), co-trimoxazole (22, 23), ofloxacin (27, 28), or ciprofloxacin (26). While the cohort studies had no clear doses and durations (22, 23), the *Brucella* clinical trials mostly investigated a standard dose of 200 mg doxycycline daily + 600 mg rifampin daily vs. 400 mg ofloxacin daily + 600 mg rifampin daily for 6 weeks (27, 28). *Chlamydia*-induced reactive arthritis studies compared a dose of 200 mg doxycycline daily vs. placebo (24, 25). Non-dental studies are further summarized in Tables 1 & 2. Periodontitis studies compared a dose of 100 mg doxycycline daily vs. placebo and usual care or usual care alone for 14 to 21 days (Table 3) (29–31).

**Table 3.** Summary of dental clinical trials eligible for inclusion in systematic review.

| Study | Country | Design | Sample size (N) | Study population | Site | Intervention | Control | Outcome |
| --- | --- | --- | --- | --- | --- | --- | --- | --- |
| **Chaves 2000** <sup>(26)</sup> | USA | Randomized clinical trial | 40; intervention:20; control:20 | Adults (30-70 years) | Periodontitis within the last 2 years | Scaling and root planing + doxycycline: loading: 200 mg the 1st day; 100 mg daily for 21 days | Scaling and root planing | No significant difference between the intervention and the control in bacterial antigens, including *P. gingivalis*, and mean bone loss |
| **Tsalikis 2014** <sup>(27)</sup> | Greece | Randomized, controlled study | 70; intervention:31; control:35 | Adults, age > 30 years with controlled type 2 diabetes | Chronic periodontitis | Doxycycline: loading: 200 mg the 1st day; 100 mg daily for 21 days | Placebo with the same instructions | No significant difference between the intervention and the control in clinical assessment (PD and CAL) and microbiological species, including *P. gingivalis* |
| **Al-Nowaiser 2014** <sup>(28)</sup> | Saudi Arabia | Multi-center, randomized, parallel, single-blind study | 68; intervention:35; control:33 | Adults (21-80 years) | Moderate to severe chronic periodontitis | Scaling and root planing + doxycycline: loading: 200 mg the 1st day; 100 mg daily for 14 days | Scaling and root planing | No significant difference between the intervention and the control in clinical assessment (PD and CAL); significant reduction in *P. gingivalis* population with doxycycline after 1 month but not after 3 and 6 months |

PD: probing depth; CAL: clinical attachment level.

### Outcomes

Relapse frequency and relapse rate were the most frequently reported outcomes in *Brucella* studies (22, 23, 26–28). *Chlamydia* studies assessed pain scores (25), joint swelling, and joint tenderness (24), while dental studies mainly reported probing depth and clinical attachment level (30, 31).

### Risk of Bias in Included Studies

The Newcastle-Ottawa quality assessment for cohort studies showed a minimum score of 4 (Figure S1). However, all the identified cohort studies got zero stars in the comparability domain and were assessed as being of low quality due to the lack of control for confounders (21–23). The clinical trials assessment indicated that the included studies also had a high risk of bias; six studies were judged to be at high risk of bias in at least one domain (26–31), while the remaining two trials were judged to have some concerns in multiple domains (24, 25). Clinical trial assessments are displayed in Figures S2 & S3.

### Meta-Analyses Results

The pooled analysis of the two clinical trials that compared the relapse risk of *Brucella* arthritis after six weeks of treatment with either 200 mg doxycycline daily + 600 mg rifampin daily or 400 mg ofloxacin daily + 600 mg rifampin daily found no significant difference in the relapse risk between the two combinations (118 participants; RR: 0.94, 95% CI: 0.2 - 4.45, I^2^ =L0%) (Figure 2). Also, the pooled analyses of the two clinical trials that compared probing depth and clinical attachment level between 100 mg doxycycline (2-3 weeks) and no antibiotic among chronic periodontitis patients found no significant difference after 3 months of follow-up (134 participants; RR: 0.16, 95% CI: -0.18 – 0.5, I^2^ =L0%; RR: 0.05, 95% CI: -0.29 – 0.4, L=L0%, respectively) (Figures S4 & S5). Additionally, no significant difference was found after 6 months of follow-up (134 participants; RR: -0.09, 95% CI: -0.34 – 0.15, I^2^ =L0%; RR: 0.06, 95% CI: -0.26 – 0.38, I^2^ =L0%, respectively) (Figures S6 & S7).

**Figure 2.**
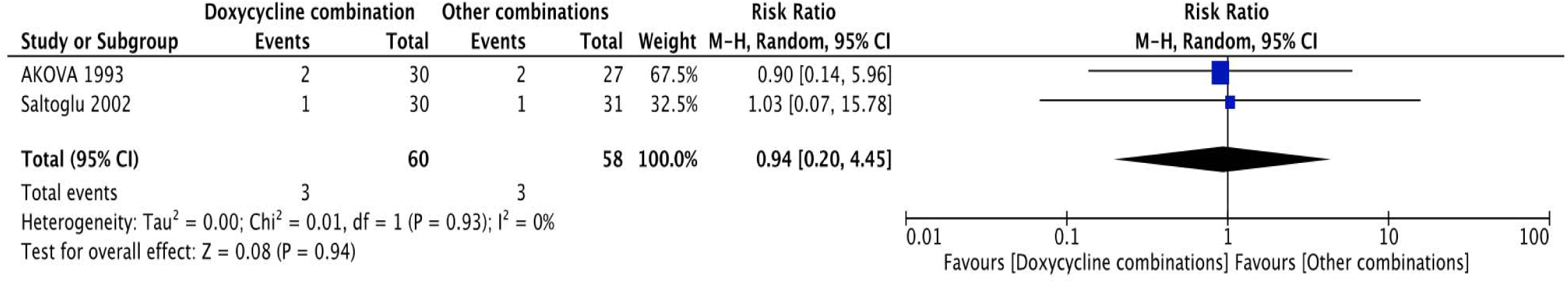
*Brucella* arthritis relapse risk after 6 weeks of treatment with doxycycline + rifampin vs. ofloxacin + rifampin.

## DISCUSSION

In this systematic review of the available literature about doxycycline-based treatment of musculoskeletal infections, we identified a high prevalence of case reports and case series, along with some cross-sectional studies. Such designs provide limited value for understanding doxycycline treatment outcomes, given the small number of patients, the heterogeneity of cases, and the limited ability to account for temporality or confounding with these designs. Furthermore, our review found that the evidence pertaining to the most common musculoskeletal infections treated with doxycycline—e.g., staphylococcal infections—was sparse. Of eight reports investigating doxycycline outcomes for medical/non-dental musculoskeletal infections, five studied *Brucella*. These studies were often limited by sample sizes and power considerations, owing in part to the rarity of these infections, yet studies of more common infections (with higher possible sample sizes) were lacking. This is a crucial gap in the literature informing doxycycline use in clinical practice. Although doxycycline is indicated and prescribed for many Gram-positive and Gram-negative causes of osteomyelitis or septic arthritis in practice (32), particularly where increased reliance on oral therapies is desired, there is a paucity of studies examining the outcomes of this practice on non-*Brucella* musculoskeletal infections.

Overall, the pooled analyses in this study found no significant differences between doxycycline and comparison groups, which might be due to the lack of real difference, the scarcity of published studies, or methodological limitations of the included studies (e.g., low statistical power and/or lack of control for confounders). The two clinical trials examining *Brucella* relapse rates shared several methodological features, including the intervention, the control, the intervention and control dose and duration, and the participant age (27, 28). Although the total follow-up period varied between these studies (6 vs. 12 months), a minimum of 6 months follow-up after treatment completion is sufficient to assess relapse (33). Likewise, the two trials included in the periodontitis treatment outcomes meta-analyses have similar methodological characteristics except for the treatment duration (2 vs. 3 weeks); and we opted to pool the estimates since both are considered short-duration treatment for chronic periodontitis. Overall, we do not expect these minor differences to affect the main findings of the meta-analyses (30, 31).

Musculoskeletal infection treatment is complex, lengthy, and costly. For instance, between 2001 and 2011, hospital costs associated with periprosthetic joint infections alone increased from $365 to $771 million (34). Many of these costs are related to the use or monitoring of intravenous antibiotics (34), which could be partially alleviated with transition to oral antibiotic therapy with doxycycline or similar agents. Addressing the gaps identified in this review may better inform clinical practice and may prove more cost-effective in the long run. Future research, including well-designed cohort studies using existing healthcare network datasets and clinical trials, may help to better understand the effectiveness and cost-effectiveness of doxycycline in the treatment of common musculoskeletal infections.

This review has some limitations. We only searched two databases (Medline and Embase); and we did not search any grey literature databases, which might limit the comprehensiveness of the review. However, the main aim of searching a grey literature database is to reduce the risk of publication bias by including studies with null or negative results (35); and since all included studies in this review reported null findings, we do not expect this to have a major effect on the study results. Additionally, we only included studies published in English.

On the other hand, this review has several strengths. Every step was completed independently by two reviewers. The combination of Medline and Embase databases is expected to have a high indexing rate of published literature (above 90% for treatment reports) (36). Additionally, we considered both clinical trials and cohort studies in this review. Also, random effect modeling was used to conduct the meta-analyses; we opted to use a conservative approach even though there was no apparent statistical heterogeneity.

In conclusion, there is a scarcity of high-quality evidence examining doxycycline treatment outcomes for musculoskeletal infections, including the most common musculoskeletal infections encountered in clinical practice. Most of the existing evidence is focused on one relatively rare cause of musculoskeletal infection (*Brucella*) and on chronic periodontal disease with bone involvement. Pooled estimates comparing *Brucella* relapse risk and periodontal disease outcomes showed no significant differences between doxycycline-based combinations and non-doxycycline-based combinations. Further research about doxycycline effectiveness and cost-effectiveness is needed to provide more robust evidence about musculoskeletal infection care.

## DISCLOSURES

### Data Availability

This research involves data from publicly available databases (Medline, Embase). Research findings are presented in detail in the manuscript, tables, and figures.

Additional information or data are available on reasonable request to the corresponding author.

