## Supplemental Figures S1-S7 for "Treatment Outcomes of Doxycycline Use for Musculoskeletal Infections: A Systematic Review and Meta-Analysis"

**Figure S1.** Newcastle-Ottawa quality assessment for eligible cohort studies

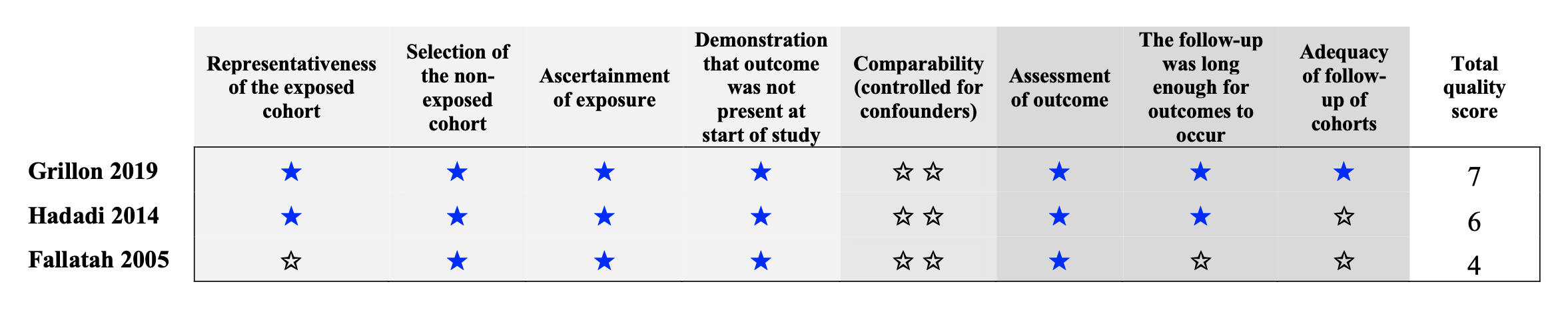

**Figure S2.** Risk of bias graph for all eligible clinical trials

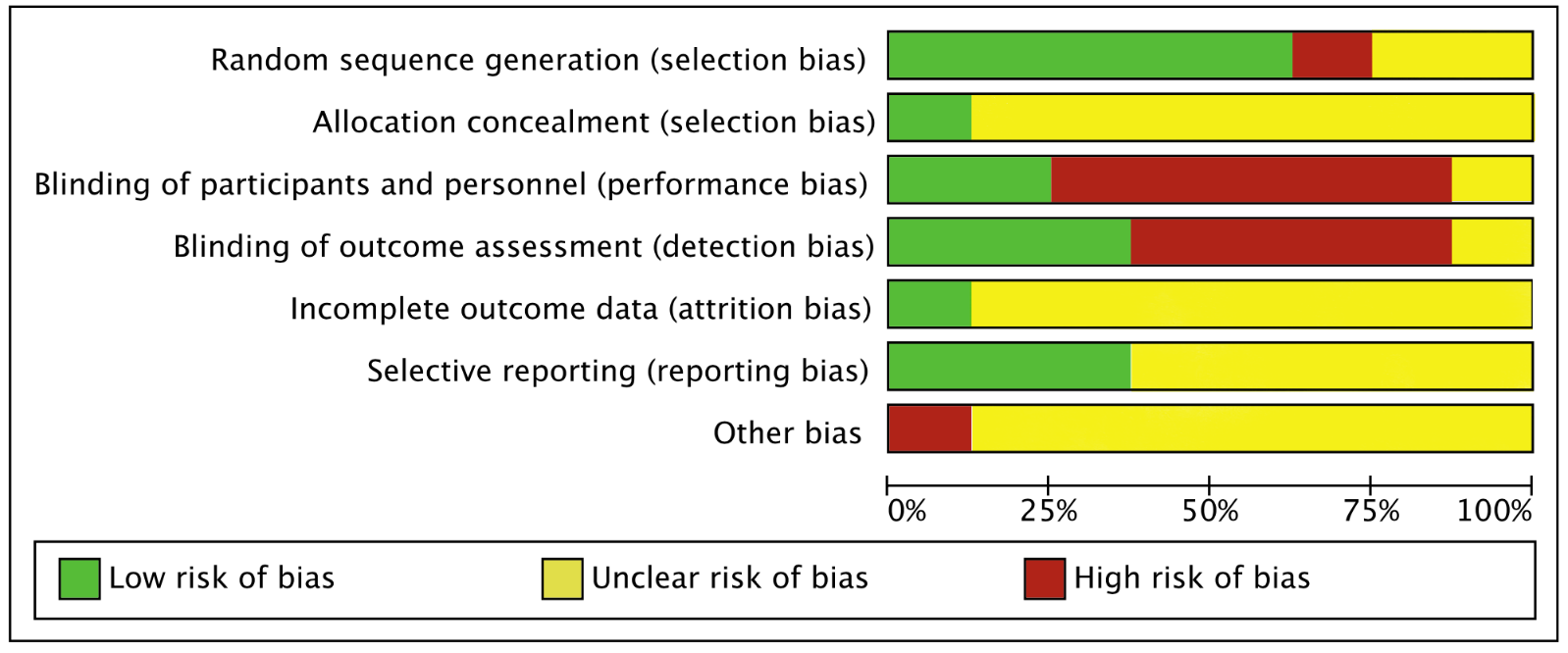

**Figure S3.** Risk of bias summary for eligible clinical trials

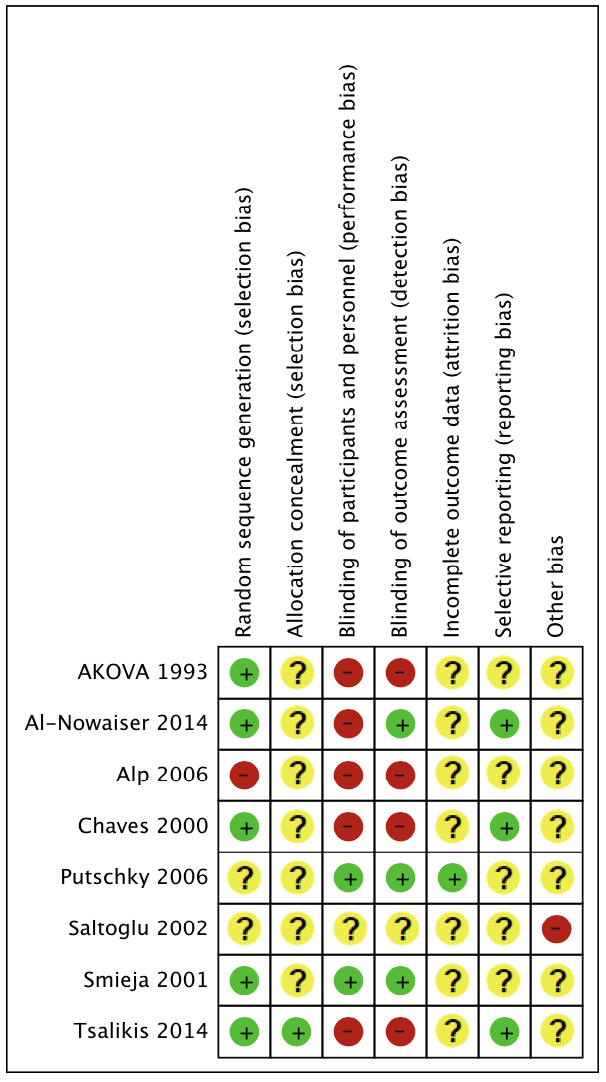

**Figure S4.** Dental outcomes for doxycycline 100 mg daily (2-3 weeks) vs. no antibiotic for chronic periodontitis, based on probing depth at 3-months follow up

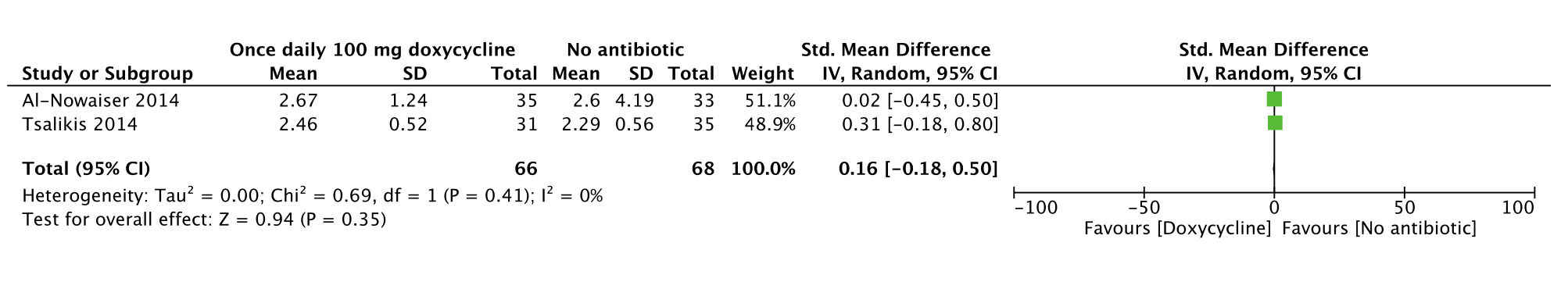

**Figure S5.** Dental outcomes for doxycycline 100 mg daily (2-3 weeks) vs. no antibiotic for chronic periodontitis, based on clinical attachment level at 3-months follow up

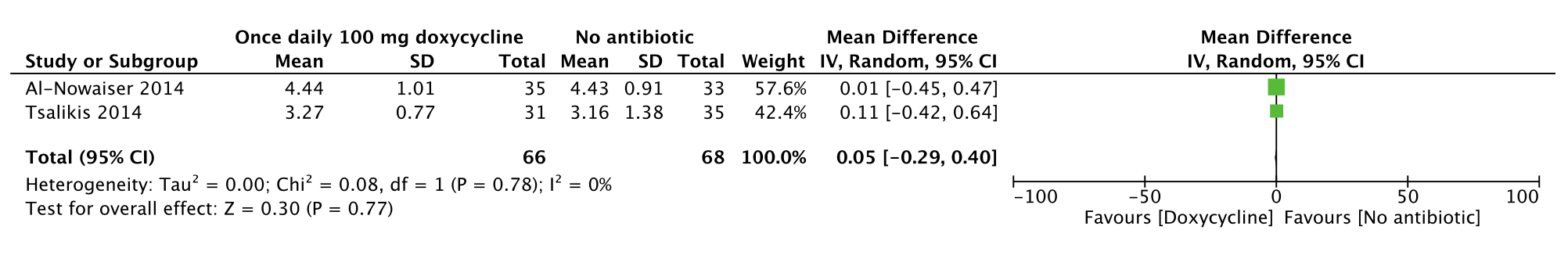

**Figure S6.** Dental outcomes for doxycycline 100 mg daily (2-3 weeks) vs. no antibiotic for chronic periodontitis, based on probing depth at 6-months follow up

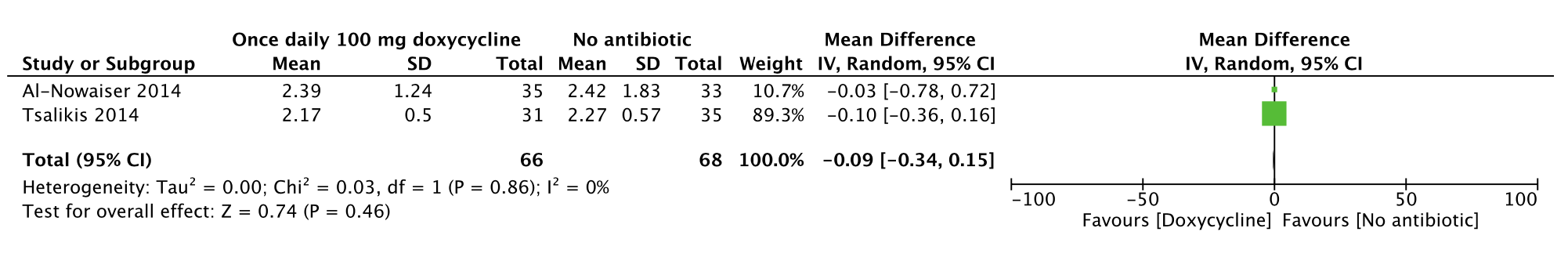

**Figure S7.** Dental outcomes for doxycycline 100 mg daily (2-3 weeks) vs. no antibiotic for chronic periodontitis, based on clinical attachment level at 6-months follow up

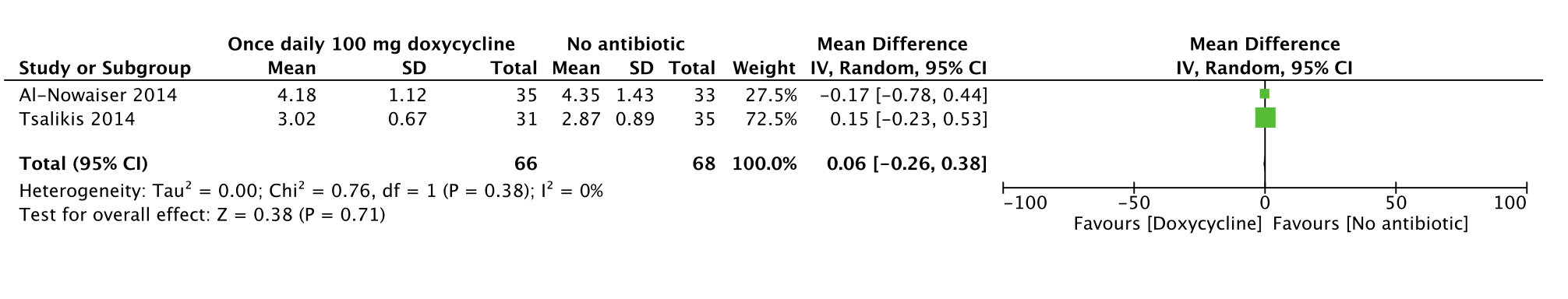
